# Influenza Vaccination and COVID19 Mortality in the USA

**DOI:** 10.1101/2020.06.24.20129817

**Authors:** Claudio Zanettini, Mohamed Omar, Wikum Dinalankara, Eddie Luidy Imada, Elizabeth Colantuoni, Giovanni Parmigiani, Luigi Marchionni

## Abstract

COVID-19 mortality rate is higher in the elderly and in those with preexisting chronic medical conditions. The elderly also suffer from increased morbidity and mortality from seasonal influenza infection, and thus annual influenza vaccination is recommended for them.

In this study, we explore a possible area-level association between influenza vaccination coverage in people aged 65 years and older and the number of deaths from COVID-19. To this end, we used COVID-19 data until June 10, 2020 together with population health data for the United States at the county level. We fit quasi-Poisson regression models using influenza vaccination coverage in the elderly population as the independent variable and the number of deaths from COVID-19 as the outcome variable. We adjusted for a wide array of potential confounding variables using both county-level generalized propensity scores for influenza vaccination rates, as well as direct adjustment.

Our results suggest that influenza vaccination coverage in the elderly population is negatively associated with mortality from COVID-19. This finding is robust to using different analysis periods, different thresholds for inclusion of counties, and a variety of methodologies for confounding adjustment.

In conclusion, our results suggest a potential protective effect of the influenza vaccine on COVID-19 mortality in the elderly population. The significant public health implications of this possibility point to an urgent need for studying the relationship between influenza vaccination and COVID-19 mortality at the individual level, to investigate both the epidemiology and any underlying biological mechanism.

## Introduction

COVID-19, a disease caused by the severe acute respiratory syndrome coronavirus 2 (SARS-CoV-2), has become a global health threat owing to its high rate of spread and mortality. By June 10, 2020, the total number of cases worldwide had reached more than 7 million with approximately 411,195 confirmed deaths^1^. The disease started in Wuhan, China in December 2019 as a zoonotic infection, but strong evidence suggests that efficient human-to-human transmission started as early as mid-December 2019^2^. Such person-to-person transmission occurs mainly through respiratory droplets with several studies reporting the possibility of spread from asymptomatic patients^3,4^.

Most symptomatic infections are mild with nearly 14% of infected individuals developing severe disease with dyspnea and hypoxia. Critical illness has been seen in only 5% of cases in the form of septic shock and respiratory and multi-organ failure^5^. In symptomatic patients, the most frequent presentation is pneumonia, manifested by fever, fatigue, dry cough, dyspnea, and pulmonary infiltration^6,7^. Other symptoms have also been reported including, but not limited to sore throat, nausea, diarrhea, myalgia, confusion, anosmia, and other taste abnormalities^8,9^. The risk of developing severe complications and mortality rates is higher in the elderly, in males, and in patients with co-morbidities, especially hypertension, diabetes, and chronic respiratory conditions including asthma and chronic obstructive pulmonary disease (COPD)^10,11^. Acute respiratory distress syndrome (ARDS) is the most common complication in people with severe illness with the incidence being higher in the older population.

Seasonal respiratory viral co-infections, most commonly influenza A and B, have been reported in COVID-19 patients^12,13^. Seasonal influenza causes distinct outbreaks every year with the attack rate varying from 10% to 20%. Similar to COVID-19, the morbidity and mortality are also higher in the older population and in patients with chronic comorbidities^14^. Thus, routine annual vaccination is recommended in these groups. While in the elderly the vaccine is of clear benefit, the benefit is even higher in those with high-risk illnesses. The absolute risk reduction from the vaccine was 2 to 4 folds higher in the elderly population with chronic underlying conditions compared to the healthy population of the same age group, even when there was a poor match between the vaccine and circulating strains^15^, and when the vaccine effectiveness rate was as low as 10%^16^. Another study found that the vaccine was effective at reducing the rate of hospitalization from pneumonia and also the rate of death in elderly people with chronic lung diseases^17^. Interestingly, Taksler et al.^18^ found an inverse relationship between influenza vaccine coverage in adults aged between 18 and 64 years and influenza-related illness in the older population (>= 65*years*) which can be explained by reduced transmission in the community.

Since there is no effective treatment or vaccine available for COVID-19 to this date, other measures have to be taken to reduce the morbidity and mortality of this disease. Public health measures like isolation, quarantine, and social distancing are being used worldwide to contain outbreaks and reduce the spread of the disease^19^. These measures are key not only to mitigate the virus transmission but also to better manage healthcare resources^20^.

A plausible conjecture is that reducing the rate of chronic and acute respiratory comorbidities in high-risk populations, could in turn lead to a reduction in the complications and deaths associated with COVID-19. Based on this, it could be hypothesized that routine influenza vaccination may be associated with lower morbidity and mortality from COVID-19 because immunized individuals will have a lower rate of respiratory complications from influenza, and thus will have a better baseline health condition to fight SARS-CoV-2 infection. On the other hand, however, increased odds of non-SARS coronaviruses infection have been recently reported among army personnel who received influenza vaccination^21^. Therefore, it is possible that influenza vaccination may increase the susceptibility to SARS-CoV2 infection, and thus be associated with increased COVID-19 morbidity, and mortality, through immune-mediated mechanisms like antibody-dependent enhancement (ADE)^22^.

Motivated by these observations, we investigate whether influenza vaccination coverage is associated with COVID-19 mortality, and, if so, in which direction. To this end, we analyze COVID-19 mortality rate and influenza vaccination coverage in the United States (US) at the county level. While an area-level association can only provide preliminary evidence, this issue requires urgent attention, and the data we collected is the most effective in shedding light on it at this time. In our analysis, we take great care to adjust for important social, economic, and health determinants, to mitigate the intrinsic limitation of observational studies based on aggregated data.

## Materials and Methods

### Social, economic, demographic, and COVID-19 variables

To evaluate the impact of influenza vaccination on COVID-19 mortality, we collected and analyzed data on vaccination coverage, COVID-19 mortality, and other important social, economic, and health variables from the United States of America (US). The cumulative number of COVID-19 cases and deaths were considered until June 10, 2020.

### Data sources

The number of COVID-19 confirmed cases and deaths were obtained from the Johns Hopkins University (JHU) Center for Systems Science and Engineering (CSSE) (https://github.com/CSSEGISandData/COVID-19/), which aggregates and combines data reported by national and state sources. Up to date statistics on COVID-19 testing for each state were obtained from the COVID Tracking Project (https://covidtracking.com/api), an organization that aggregates metrics from different sources regarding the COVID-19 epidemic in the United States. All these statistics include cases in which the presence of SARS-CoV-2 was laboratory-confirmed, and others in which it was presumed and considered the probable cause of death^23^. For Rhode Island, the JHU repository reported deaths only at the state level. Thus, the number of deaths in each county was obtained using the New York Times COVID-19 data repository (https://github.com/nytimes/covid-19-data). Information on influenza vaccination coverage in people aged 65 and older, medical conditions, and tobacco use (year 2017) were obtained from the Center of Medicare Disparity Office of Minority Health (https://data.cms.gov/mapping-medicare-disparities). Demographic data on household composition, gender, race, age, and poverty levels (year 2018) in each US county were retrieved from the Census Bureau (https://data.census.gov/cedsci/table?q=United%20States. The number of hospital beds per county (year 2020) was obtained from the Homeland Infrastructure Foundation (https://hifld-geoplatform.opendata.arcgis.com/datasets/hospitals/data?page=18). Finally, information on the emissions of particulate matter (*PM*_*2*.*5*_, year 2016) was originally reported by the Atmospheric Composition Analysis Group (http://fizz.phys.dal.ca/~atmos/martin/?page_id=140#V4.NA.02.MAPLE) and obtained and pre-processed from Wu et. al^24^.

### Data and software availability

All data used in this study, along with detailed information about data sources, are available in the form of the R package `covid19census`, freely distributed from GitHub (https://github.com/c1au6i0/covid19census). All scripts and code used to pre-process and analyze the data are available from https://github.com/c1au6i0/covid19_influenza. All data pre-processing and statistical analyses in this study were performed using the R programming language^25^, and libraries from the `tidyverse` suite^26,27^.

### Inclusion criteria

Information was available for a total of 3243 counties across the 50 states and the Washington D.C. district, between January 22, 2020 and June 10, 2020. For a total of 1219 COVID-19 deaths, the county of the deceased was unknown. Those deaths were omitted from the analysis since also information regarding the variable of interest and the confounders were not available. As of the last day of observation, a total of 206 counties did not report any confirmed cases. To limit the impact of limited SARS-CoV-2 community circulation and exposure on COVID-19 mortality, only counties with at least 10 COVID-19 cases were included in the models (n=2034, see Supplementary Figure S1). This also allowed to calculate and include the duration of the outbreak (in the form of the number of days elapsed since the first reported COVID-19 case) among the confounding variables. For all other variables, we used the latest available year information, as summarized below.

### Potential Confounders

Overall, the initial set of variables related to population demographics, clinical and territory characteristics was large (≈300). In our analyses, therefore, we selected representative variables from each important category, and avoided highly correlated pairs. Specifically, the following variables were included in our models:

1. <u>Household-related variables</u>: a) % of family households, b) % of families with a single parent, and c) % of households with access to internet. Note that a family is defined by the US Census as consisting of “a householder and one or more other people living in the same household who are related to the householder by birth, marriage, or adoption”.
2. <u>Socioeconomic- and healthcare-related variables</u>: a) median income, b) hospital beds per person, c) number of COVID-19 tests (at the state level), d) number of days since the first case in that county.
3. <u>Education-related variables</u>: (a) % of people with a bachelor degree.
4. <u>Race-related variables (% of people in the population of the following races)</u>: a) Asian, b) Black or African-American, c) Latino or Hispanic, d) Native American, e) Caucasian or White, f) other races, g) belonging to two or more races.
5. <u>Demographic variables</u>: a) % of people 65-year of age or older, b) median age, c) sex ratio (males per 100 females), and child dependency ratio (ratio between population under 18 years and population 18-to-64).
6. <u>Medical conditions or diseases (% of people 65-year of age or older with)</u>: a) dementia, b) asthma, c) atrial fibrillation, breast cancer, e) colorectal cancer, f) lung cancer, g) obstructive pulmonary disease, h) chronic kidney disease, i) depression, j) diabetes, k) heart failure, l) hypertension, m) ischemic heart disease, n) obesity, o) rheumatoid arthritis p) stroke and transient ischemic attack, and q) tobacco use.
7. <u>Environmental variables</u>: a) fine particulate matter (*µg*/*m*^2^, average of years 2000 to 2016), b) winter and summer temperature, and c) winter and summer humidity.

### Statistical Modeling

#### Generalized linear model

We are interested in estimating the change in the COVID-19 mortality rate associated with a change in the county-level influenza vaccination coverage. To this end, we used a quasi-Poisson regression with the number of deaths from COVID-19 as the response variable, the influenza vaccination coverage in people 65-year of age and older as an independent variable, and the population size of each area as offset. We calculated the mortality rate ratio (MRR) using the the R package `oddratios`^28^. The MRR represents the ratio of COVID-19 mortality corresponding to an increase of 10% in the influenza vaccination coverage. We chose population mortality, in contrast to mortality among the infected population, to remove the effect of inconsistent COVID-19 testing policies across areas. We controlled for potential confounding both via generalized propensity scores and direct adjustment, as described next.

#### Confounding adjustment with generalized propensity score

We estimated a generalized propensity score (PS) model for county-level rates of vaccination over 65-year of age by regressing the logit of influenza vaccination coverage on selected confounding variables (see “Potential Confounders” above). The propensity score is a balancing factor that allows to correct for the potentially unequal distribution of explanatory and confounding variables across levels of exposure^29–31^. In our main analyses, we stratified the propensity score into quintiles, and then used these as a factor in the quasi-Poisson regression analysis. We performed this analysis with and without state-specific fixed effects. Additionally, we employed a Poisson mixed model in which the state was included as random factor to control for state differences. We also used the PS quintiles to perform a fully stratified analysis. Lastly, we repeated the analysis by including the propensity score as a linear term (see secondary analyses).

#### Direct confounding adjustment

As an alternative, we also modeled the effect of immunization coverage on COVID-19 mortality by directly adjusting for confounding variables via linear terms in the quasi-Poisson regression. To select a parsimonious number of potential confounders, we clustered candidate variables based on their correlation with each other and selected a single variable from each cluster (see Supplementary Figure S3). Specifically, we selected: a) % of family households, b) rate of hospital beds, c) % of people with asthma, d) % of people with breast cancer, e) % of people with lung cancer, f) % of people with COPD, g) % of people with hypertension, h) median income, i) summer temperature, j) summer humidity, k) winter temperature, l) winter humidity, m) % of people >= 65 years old, n) sex ratio, and o) days passed since the first case.

#### Secondary analyses: Dependency on counties with highest and lowest infection rates

Counties with extremely low or high numbers of confirmed cases (and deaths) might have undue leverage on statistical estimates. To explore sensitivity of our results, we repeated the analysis by requiring a larger minimum number of confirmed cases for inclusion in the model (ranging from 10 to 100 in increments of 10). Also, since New York State has the highest number of confirmed cases (and deaths) from COVID-19 in the US, we repeated the analysis after removing all 53 New York State counties. Finally, since we noticed that for 46 counties, no deaths were reported despite the large number of cases (*i*.*e*., exceeding 100), we hypothesized that different criteria could have been used to record COVID-19 related deaths in these counties. We therefore repeated our analysis including only the counties with at least one death.

#### Secondary analyses: Stability over time

Our analyses are somewhat arbitrarily based on the time window ending on June 10. As information on COVID-19 continues to accrue results may evolve. To assess stability across time of our estimated effects, we repeated the analyses with the data available at different points over approximately two months.

#### Independent Verification: Exposure to air pollution

Levels of airborne fine particulate matter (*PM*_*2*.*5*_) have been reported to contribute to COVID-19 mortality^24^, based on analyses sharing much of the same data structure and methodology with what done here. To provide an independent verification of our data collection and analysis pipeline, we repeated our analysis using chronic exposure to fine particulate matter (*PM*_*2*.*5*_) as an alternative exposure, and including influenza vaccination coverage among the possible confounding variables. We were able to reproduce the reported effect of *PM*_*2*.*5*_ exposure on COVID-19 death rate.

## Results

### Social, economic, demographic population characteristics and COVID-19 outcomes in the US

A total of 2034 counties were included in the analysis. The median influenza vaccination coverage in people ≥ 65-year of age is 45%. The COVID-19 mortality rate did not differ substantially between counties where vaccination coverage was above the median (20.5 ± 52 sd), compared to those where it was below (17.3 ± 31.7 sd). Counties with lower influenza-vaccination coverage tend to have a lower level of education and income, and to have a higher percentage of Black and Latino populations. In contrast, counties with higher vaccination coverage tend to be more affluent, and to have an higher percentage of white population (see Table 1). Other important health, social, and demographic factors do not appear to be differently distributed between counties with different vaccination coverage.

**Table 1.**
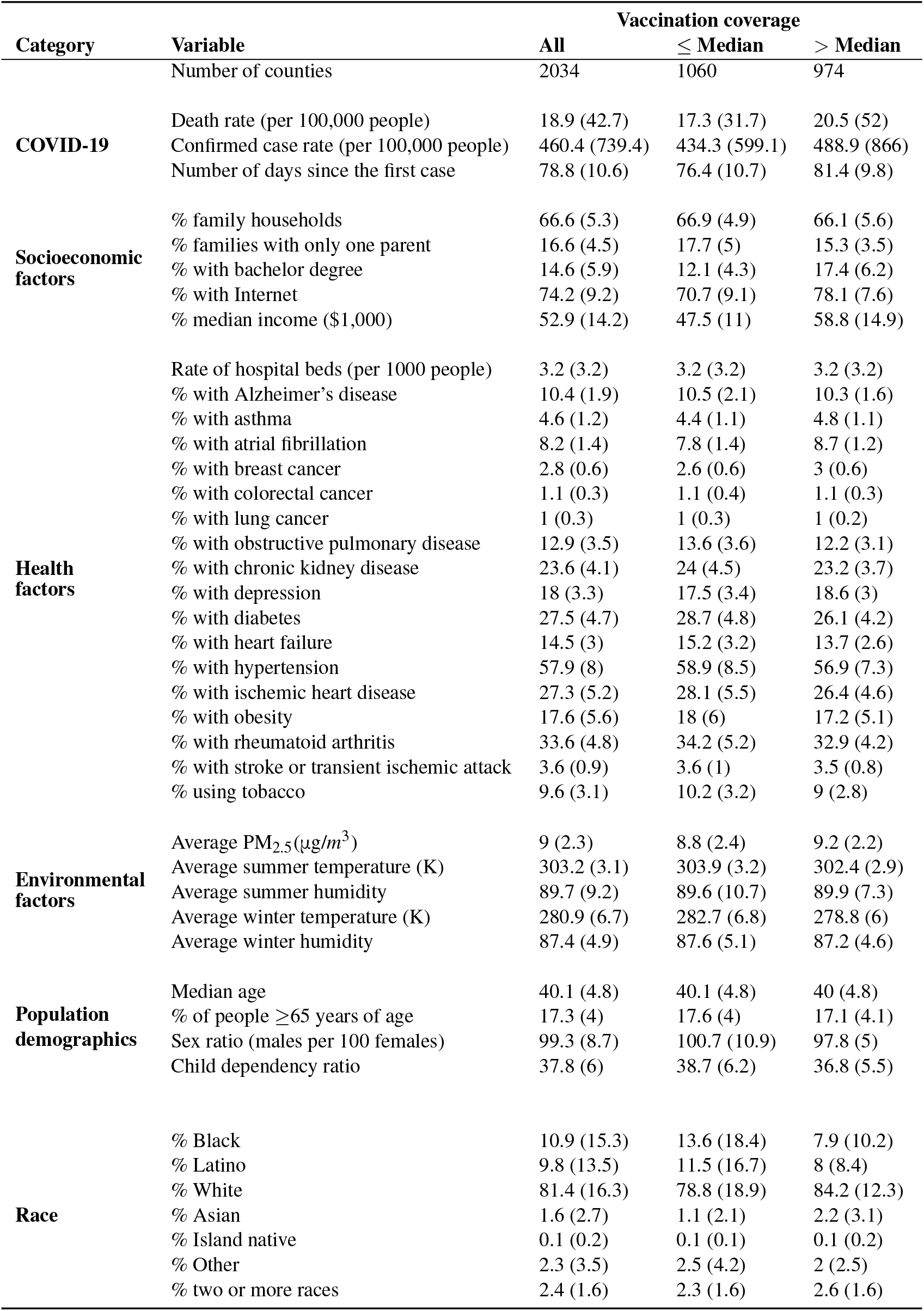
County-level data for the United States. Counties, population characteristics, and other important social, economic, and health related metrics used in the analyses. Data are presented overall and stratified by counties with vaccination coverage below and above the median of 45%. In the Table, % with chronic conditions refers to those aged 65-year and older only. Standard deviations are in parenthesis.

### Influenza vaccination and COVID19 mortality

#### Primary analyses

Several variables had a significant effect on influenza vaccination coverage, including demographic factors like the race and education level, together with a number of chronic health conditions like diabetes and hypertension (see Supplementary Table S1 for details). We therefore adjusted our model using the propensity score of being vaccinated as factor after stratification into quintiles. Such generalized linear model produced a significant negative coefficient for influenza vaccination coverage (*estimate* = −3.29, *t* = −3.01, *p* < 0.01, *d f* = 2028, see Table 2 and Supplementary Table S2). Thus, after controlling for confounding variables that could affect both the proportion of vaccinated individuals and mortality in an area (Table 1), our analyses suggest that higher influenza vaccination coverage is associated with lower COVID-19 mortality rates. Specifically, for every 10% increase in influenza vaccination coverage, there is a 28% decrease in the rate of mortality from COVID-19 (*MRR* = 0.72, 95%*CI* = 0.58−0.89, see Figure 1). Such protective effect, however, appears to be non-linear, being stronger in the counties where the vaccination coverage is higher. Indeed, when we analyzed the link between vaccination coverage and COVID-19 mortality within each US county group as defined by the propensity score quintiles, the effect was stronger in the two upper quintiles strata (see Supplementary Table S6).

**Table 2.** Effects of vaccination coverage in the US using different adjustments. Results of the different regressions performed in our study. Effects are reported for all models fitted to capture the link between influenza vaccination coverage and COVID mortality in the US. Each line represents a distinct model type or adjustment.“Vaccination” refers to county-level influenza vaccination coverage in people >= 65 years. “PS” refers to vaccination propensity score. All the models are quasi-Poisson with t-statistic, with exception of the mixed effects one which is a Poisson model with Z-score statistic.

| Vaccination effect | estimate | std.error | statistics | p.value |
| --- | --- | --- | --- | --- |
| Corrected by PS quintiles | -3.29 | 1.10 | -3.01 | 0.00 |
| Corrected by continuous PS | -5.65 | 1.21 | -4.67 | 0.00 |
| Corrected by PS quintiles and US state (fixed effects) | -5.64 | 0.62 | -9.17 | < 0.00 |
| Corrected by PS quintiles and US state (mixed effects) | -5.64 | 0.08 | -69.01 | 0.00 |

**Figure 1.**
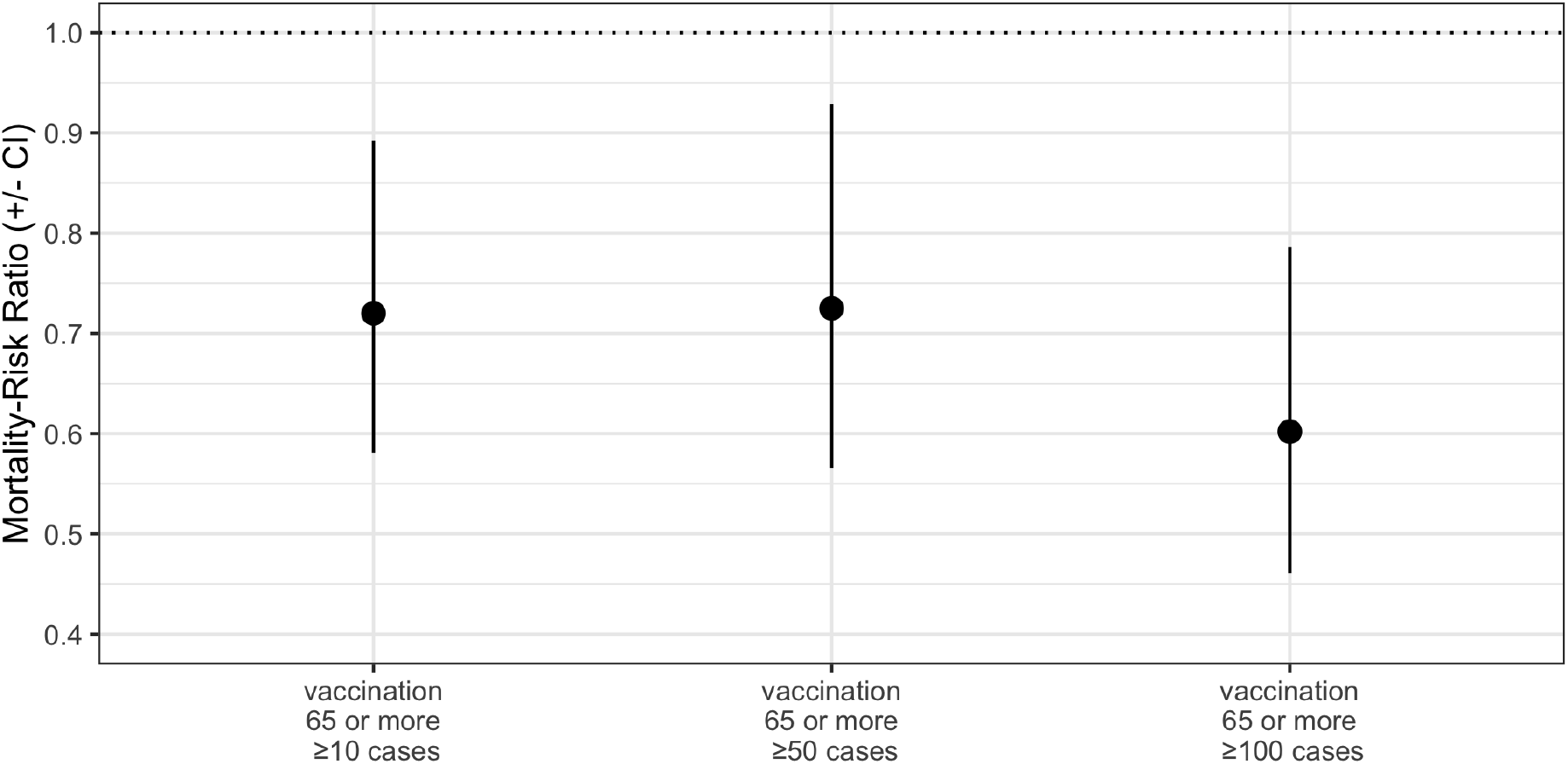
Mortality rate ratio (MRR) and its 95% CI. The MMR for influenza vaccination is the percentage change in the death rate from COVID-19 associated with a 10% increase in vaccination coverage. Results shown are for the primary analysis performed on all counties with more than 10 cases, and for the secondary analyses where distinct thresholds for inclusion where used (respectively, more than 50 and 100 confirmed cases).

#### Secondary analyses

We further evaluated the robustness of this effect using alternative approaches. Including the propensity score as a linear term in the quasi-Poisson model yielded similar results (*estimate* = −5.65, *t* =− 4.66, *p* < 0.01, *d f* = 2031, see Supplementary Table S3). Furthermore, we also controlled for potential confounders arising from differences in healthcare access and testing policies at the state level using a fixed effects model (*estimate* = −5.64, *t* = −9.16, *p* < 2*e*−16, *d f* = 2027, see Table S4) and a mixed effects model (*estimate* = −5.64, *t* = −69.00, *p* < 2*e* −16, *d f* = 2027, see Table S5). The effect of influenza vaccination coverage on COVID-19 mortality remained stable over time, indicating that, at this stage, the accumulation of new data is unlikely to strongly affect the results of our analysis (Figure S2). We also adjusted for the confounding variables directly as described in the methods section. The results showed that the effect of influenza vaccination remained significant (*estimate* = −4.67, *t* = −6.07, *p* < 0.01, *d f* = 2017; see Supplementary Table S8).

The effect of vaccination coverage on COVID-19 mortality in the US was not significant after removing New York state from the analysis (*estimate* = − 0.26, *t* = − 0.406, *p* = 0.69, *d f* = 1975) but it was significant when adjusting for propensity score as a continuous variable (*estimate* = −2.14, *t* = −2.96, *p* = 0.00, *d f* = 1975, see Supplementary Table S7).

By repeating our analysis including only the counties with at least one death, the effect of vaccination remained significant indicating that it is not affected by the counties with zero mortality (*estimate* = −3.10, *t* = − 2.51, *p* = 0.01, *d f* = 1556). Finally, increased air levels of particulate matter (*PM*_*2*.*5*_) was found to be associated with a higher number of deaths from COVID-19 (*estimate* = 0.09, *t* = 3.33, *p* < 0.01, *d f* = 2028), as previously reported^24^. Overall, the results from all these analyses were consistent, further confirming that the link between influenza vaccination and reduced mortality from COVID-19 is robust.

## Discussion

The main motivation for our study was to gather preliminary evidence about a possible connection between influenza vaccination in the elderly population and the risk of mortality from COVID-19. On one hand, seasonal influenza infection is associated with the development of several respiratory complications, especially in the elderly^14^. On the other hand, a recent report points to increased odds of non-SARS coronaviruses infections among army personnel who received influenza vaccination^21^. This observation was attributed by the authors to the possibility that vaccinated individuals may lack the non-specific immunity acquired by natural influenza infection, which would protect against infection by other viruses^32,33^.

Public county-level data in the U.S.A. provide a platform to begin exploring this issue. We quantified the county-level association between influenza vaccine coverage in people older than 65 years and COVID-19-related deaths in these data. We focused on people aged 65 years and older since this age group is more susceptible to influenza complications and thus annual influenza vaccination is recommended for them. This age group also has a higher mortality rate and a greater risk for developing severe complications from COVID-19, compared to the younger population^10^. In our analyses, we controlled for a wide array of potential confounding variables, including population density, social and economic variables, education, chronic medical conditions, and other important demographic and environmental factors.

Our results suggest a reduction in COVID-19 mortality associated with higher influenza vaccination rates in the elderly population. Specifically, we found that overall, a 10% increase in vaccination coverage was associated on average with a statistically significant 28% decrease in the COVID-19 death rate. Our findings suggest that influenza vaccination can play a protective role in COVID-19, and that additional confirmatory studies at the individual level are urgently needed.

This conclusion is robust to several variations of our analytical techniques. We varied the time frame considered, and our threshold for inclusion of counties with low exposure to SARS-CoV-2. We also investigated systematically several variations in the methodology used for adjusting for potential confounding factors.

Among the analyses we considered is complete stratification by quintiles of the propensity score. This analysis provides five separate effect estimates. When these are averaged, the results are consistent with the analysis of Supplementary Table S6. However, the effects within the strata differ, with groups of counties with higher generalized propensity scores manifesting higher effects. Our propensity score reflect a large number of potential confounders, many of which have an important effect. Thus, further analyses are needed to shed light on the potential sources of this heterogeneity. Generally speaking, however, our analysis considers COVID-19 mortality per inhabitant, and thus it is possible that higher county-level vaccination rates may by themselves provide control of mortality via herd immunity.

Our area-level association is consistent with a protective effect, which can be explained by the vaccine’s effect of reducing the rate of hospitalization and severe pulmonary complications in the elderly. Evidence suggests that the vaccine benefit is greater in people with high-risk respiratory conditions like chronic obstructive pulmonary disease^34^ and asthma^35^. Furthermore, influenza vaccination was found to have a protective effect against cardiovascular events^36^, confirming previous findings that linked influenza infection to increased rate of hospitalization and death from myocardial infarction^37,38^.

A complementary explanation for a putative protective effect is that unvaccinated individuals are at risk of persistent viral infections, leading to a decline in T-cell diversity which in turn impairs the immune response against other pathogens including SARS-CoV-2^39,40^. Influenza vaccination, on the other hand, does not induce a strong, virus-specific CD8 T-cell immune response, as seen with natural infection^41,42^, and while this is considered a major disadvantage of inactivated influenza vaccines, it may be beneficial in clearing SARS-CoV-2 infection, as vaccinated individuals will have more T-cell diversity (and thus better chances to fight other viruses) compared with those with natural influenza infection.

Importantly, the influenza virus has been shown to induce apoptosis and impair the cytotoxic effect of natural killer (NK) cells^43–45^, ultimately impairing the host immune defense mechanisms against other pathogens, including potentially SARS-CoV-2, especially in the acute phase of the disease. Additionally, consistently unvaccinated individuals are more likely to have a higher proportion of influenza-specific resident memory T-cells (*T*_*RM*_) in their lungs, which are highly proliferative and highly productive of inflammatory cytokines^46,47^. This, in turn, might be associated with the exaggerated inflammatory response and severe Acute Respiratory Distress Syndrome (ARDS) seen in some COVID-19 patients^48^. Furthermore, the Influenza A virus has been recently shown to up-regulate ACE2 receptors in the lung alveolar cells^49^, suggesting that a recent influenza infection can potentially lead to more severe pulmonary complications from COVID-19, in light of the fact that these same receptors are used by SARS-CoV-2 for cellular entry.

We regard ours as an urgent, but preliminary, study and acknowledge several limitations. Important potentially confounding variables include the socioeconomic levels, quality of healthcare, and the lack of uniform death reporting approaches. While our covariates allow to control for some of these confounding factors, more systematic and accurate data collection approaches would improve the reliability of analysis such as ours. An important limitation is that we could only control for the number of tests performed at the state level, since testing data were not available for each county. Furthermore, testing availability and recommendations, especially in the initial phase of the pandemic, varied between states. We tried to mitigate this issue by using the total population in the county as the offset in our models, rather than the total number of confirmed cases.

Since the number of performed tests can have an important impact on the number of reported cases, we decided not to explore the effect of influenza vaccination on SARS-CoV-2 reported infection rates. We were also unable to stratify and analyze COVID-19 death rate by age group, because this information was not available. Similarly, in our study, we could not take into account the variability in vaccine formulation and in vaccine efficacy, which depends on the predominant circulating strains, because this kind of data was not reported.

Based on the information currently available, it is unclear whether the observed negative association with COVID-19 mortality could also be observed also if we considered natural influenza infection as the exposure, and whether vaccinations against other respiratory pathogens or other diseases can confer any protection. In this regard, it was recently proposed that live attenuated vaccines, by stimulating the innate immunity, could provide transient protection against COVID-19^50^. It is also important to underscore that COVID-19 may be accompanied by innate immune response suppression^51^, therefore further suggesting that vaccination could also be beneficial, by boosting innate immunity.

Finally, an important potential confounder not captured in our dataset concerns the use of other drugs. Recent studies have proposed a potential effect of some drugs including statins and anti-hypertensives like ACEIs/ARBs on COVID-19 mortality in the elderly population^52^; however, these potential effects are still not confirmed and need further investigation.

Although many important confounding variables have been taken into account in our analyses, there is still a strong need to go beyond aggregated data to perform analyses at the individual level. These studies would provide much more robust evidence of a possible protective effect of influenza vaccination on COVID-19 mortality, and also help determine the underlying biological mechanisms.

If an effect consistent with our analysis was confirmed in individual level analyses, it would justify a more aggressive and targeted influenza vaccination policy in the elderly population and their caregivers, to reduce the rates of hospitalization, severe respiratory complications, and deaths from COVID-19. This would be of paramount importance given the paucity of effective treatment or vaccine for COVID-19 and the extreme pressure on healthcare systems.

## Data Availability

All data used in this study, along with detailed information about the data sources, are available in the form of the R package covid19census, freely distributed from GitHub.
All scripts and codes used to pre-process and analyze data are also available On GitHub using the links below.

https://github.com/c1au6i0/covid19census

https://github.com/c1au6i0/covid19_influenza

## Acknowledgements

We thank Drs. Francesca Dominici and Franco Solerio for their comments and suggestions that helped improve the manuscript. This publication was made possible through support from the NIH-NCI grants P30CA006973 (L.M.).

## Author contributions statement

L.M. conceived the idea of the study. CZ wrote the code used to aggregate data and perform analysis. G.P. conducted the first analysis of the data and identified the appropriate statistical models to use. E.C. independently validated the analysis. C.Z., M.O., W.D., E.L.I., G.P., E.C., L.M. all contributed to the analyses, interpreted the results, and edited the draft. All authors reviewed the manuscript.

**Figure S1.**
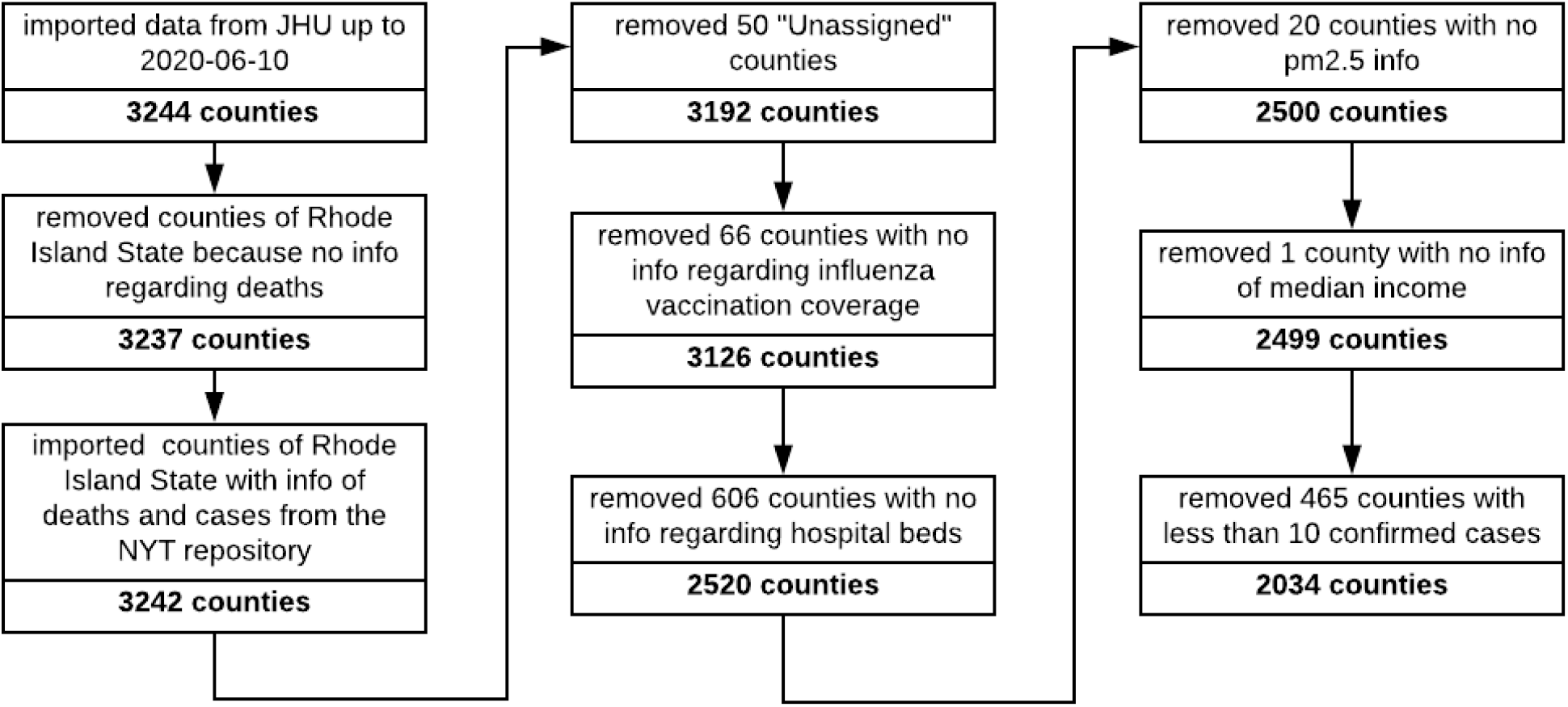
Flowchart illustrating the steps of data collection and filtering. The initial dataset comprised 3244 counties. The COVID-19 data up to June 10, 2020 were imported using the JHU repository. Since this repository does not report Rhode Island COVID-19-related deaths at the county level, that information was obtained from the NYT repository instead. Counties with missing information regarding the variable of interest or the confounders as well as those with less then 10 cases were not included in the analysis. The final dataset used for the main analysis comprises 2034 counties.

**Figure S2.**
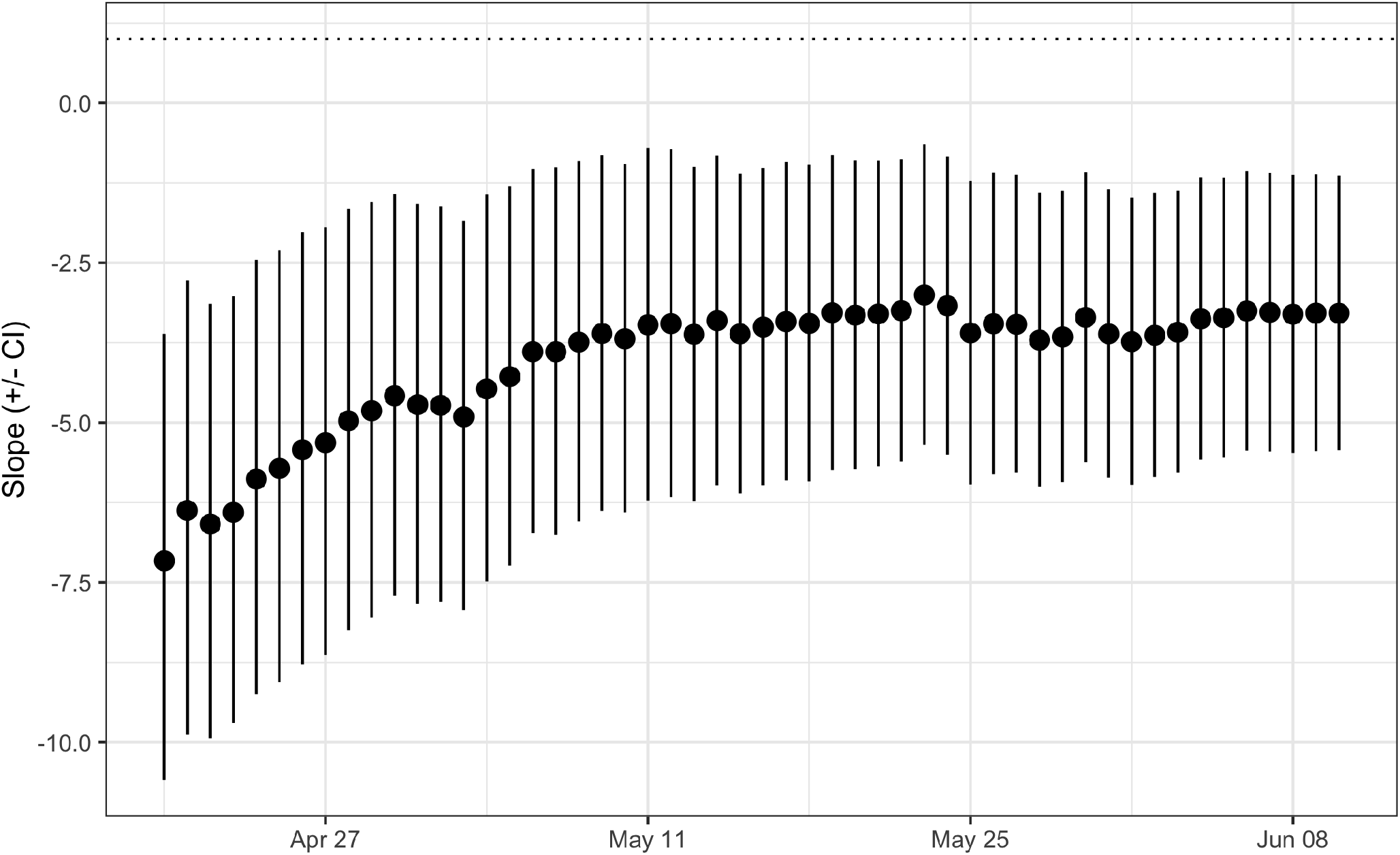
Influenza vaccination effect on COVID-19 mortality in the US using different dates as the endpoint of the analysis. Shown is the coefficients and 95% CI of the influenza vaccination coverage obtained in the quasi-Poisson regression analysis as a function of date.

**Figure S3.**
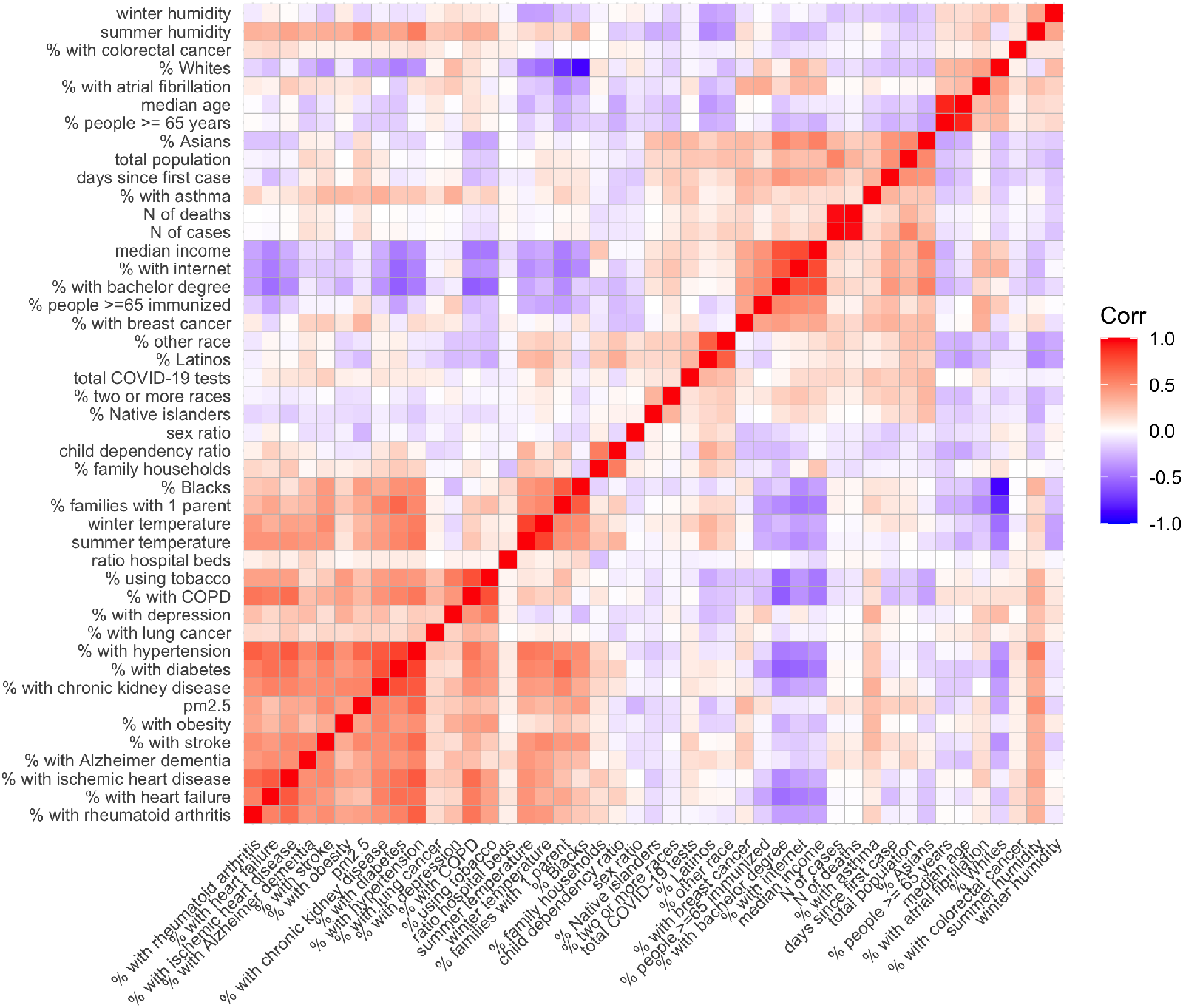
Correlation matrix of the selected confounding variables in the U.S data. Highly correlated variables are clustered together. From each cluster, we chose a single variable to be used as a covariate in the quasi-Poisson model in the direct adjustment approach.

**Table S1.** Propensity of influenza vaccination. The propensity score of vaccination in people >= 65 years based on the confounding variables. The propensity score was calculated by regressing the logit of influenza vaccination coverage on the selected confounding variables.

| Variable | estimate | std.error | t-statistic | p.value |
| --- | --- | --- | --- | --- |
| (Intercept) | -0.04 | 1.15 | -0.03 | 0.98 |
| Family households | 0.62 | 0.21 | 2.99 | 0.00 |
| Families with only one parent | -0.08 | 0.31 | -0.25 | 0.80 |
| With bachelor degree | 1.63 | 0.25 | 6.58 | 0.00 |
| With Internet | -0.00 | 0.13 | -0.02 | 0.98 |
| Ratio of hospital beds (per 100,000 people) | 3.92 | 1.97 | 1.99 | 0.05 |
| Alzheimer's disease | -1.25 | 0.47 | -2.68 | 0.01 |
| Asthma | 2.39 | 0.69 | 3.46 | 0.00 |
| Atrial fibrillation | 2.04 | 0.65 | 3.12 | 0.00 |
| Breast cancer | 4.60 | 1.29 | 3.56 | 0.00 |
| Colorectal cancer | -4.08 | 1.86 | -2.19 | 0.03 |
| Lung cancer | 1.99 | 2.49 | 0.80 | 0.43 |
| Chronic obstructive pulmonary disease | -1.50 | 0.37 | -4.04 | 0.00 |
| Chronic kidney disease | 0.83 | 0.26 | 3.22 | 0.00 |
| Depression | 1.28 | 0.30 | 4.30 | 0.00 |
| Diabetes | -1.40 | 0.32 | -4.41 | 0.00 |
| Heart failure | -0.72 | 0.34 | -2.12 | 0.04 |
| Hypertension | 1.47 | 0.22 | 6.75 | 0.00 |
| Ischemic heart disease | -0.07 | 0.22 | -0.34 | 0.74 |
| Obesity | -0.16 | 0.14 | -1.16 | 0.25 |
| Rheumatoid arthritis | -0.79 | 0.20 | -3.92 | 0.00 |
| Stroke | 2.73 | 1.00 | 2.73 | 0.01 |
| Tobacco use | -0.57 | 0.38 | -1.51 | 0.13 |
| Median income | 0.00 | 0.00 | 0.34 | 0.73 |
| pm2.5 | 0.01 | 0.00 | 3.06 | 0.00 |
| Summer temperature | 0.01 | 0.00 | 2.69 | 0.01 |
| Summer humidity | 0.00 | 0.00 | 2.37 | 0.02 |
| Winter temperature | -0.02 | 0.00 | -7.17 | 0.00 |
| Winter humidity | -0.00 | 0.00 | -1.62 | 0.11 |
| Age >=65 | 0.76 | 0.50 | 1.52 | 0.13 |
| Median age | -0.01 | 0.00 | -2.71 | 0.01 |
| Sex ratio | -0.00 | 0.00 | -4.03 | 0.00 |
| Child dependency | -0.01 | 0.00 | -4.80 | 0.00 |
| Black race | -0.62 | 0.16 | -3.90 | 0.00 |
| Latino race | -0.10 | 0.10 | -0.99 | 0.32 |
| White race | -0.10 | 0.17 | -0.60 | 0.55 |
| Asian race | -1.11 | 0.37 | -3.03 | 0.00 |
| Island native race | 6.01 | 4.46 | 1.35 | 0.18 |
| Other race | -0.10 | 0.30 | -0.35 | 0.73 |
| Two or more races | 0.68 | 0.54 | 1.27 | 0.21 |
| Total COVID-19 tests | -0.00 | 0.00 | -1.04 | 0.30 |
| Days since the first case | 0.00 | 0.00 | 2.86 | 0.00 |

**Table S2.** Effects of vaccination coverage in the US stratifying the propensity score into quintiles. Results of the quasi-Poisson regression model using the number of deaths from COVID-19 as the outcome variable, influenza vaccination coverage as the independent variable, and adjusting for propensity score quintiles. Total population was used as offset to the model.

| Variable | estimate | std.error | t-statistic | p.value |
| --- | --- | --- | --- | --- |
| Intercept | -7.46 | 0.45 | -16.61 | 0.00 |
| Influenza vaccination in people $\geq 65$ years | -3.29 | 1.10 | -3.01 | 0.00 |
| 2nd quintile | 0.50 | 0.33 | 1.53 | 0.13 |
| 3rd quintile | 0.34 | 0.33 | 1.04 | 0.30 |
| 4th quintile | 0.40 | 0.34 | 1.19 | 0.23 |
| 5th quintile | 1.76 | 0.34 | 5.19 | 0.00 |

**Table S3.** Effects of vaccination coverage in the US adjusting for the propensity score as a continuous variable. Results of the quasi-Poisson regression model using the number of deaths from COVID-19 as the outcome variable, influenza vaccination coverage as the independent variable, and adjusting for propensity score as continuous variable. Total population was used as offset.

| Variable | estimate | std.error | t-statistic | p.value |
| --- | --- | --- | --- | --- |
| Intercept | -5.05 | 0.58 | -8.63 | < 0.00 |
| Influenza vaccination in people ≥ 65 years | -5.65 | 1.21 | -4.66 | 0.00 |
| Propensity Score | 3.65 | 0.38 | 9.57 | < 0.00 |

**Table S4.** Effects of vaccination coverage in the US using a fixed effect model. Results of the quasi-Poisson regression model using the number of deaths from COVID-19 as the outcome variable, influenza vaccination coverage as the independent variable, adjusting for propensity score quintiles, and controlling for State differences using fixed effect model.

| Variable | estimate | std.error | t-statistic | p.value |
| --- | --- | --- | --- | --- |
| Intercept | -6.84 | 0.36 | -18.97 | < 0.00 |
| Influenza vaccination in people ≥ 65 years | -5.64 | 0.62 | -9.17 | < 0.00 |
| 2nd quintile | 0.33 | 0.16 | 2.07 | 0.04 |
| 3rd quintile | 0.23 | 0.16 | 1.41 | 0.16 |
| 4th quintile | 0.01 | 0.17 | 0.037 | 0.97 |
| 5th quintile | 0.93 | 0.18 | 5.18 | 0.00 |

**Table S5.** Effects of vaccination coverage in the US using a mixed effect model. Results of the Poisson regression using the number of deaths from COVID-19 as the outcome variable, influenza vaccination coverage as the independent variable, adjusting for propensity score quintiles, and controlling for State differences by including State as a random factor.

| Variable | estimate | std.error | z-value | p.value |
| --- | --- | --- | --- | --- |
| Intercept | -6.52 | 0.15 | -42.36 | 0.00 |
| Influenza vaccination in people ≥ 65 years | -5.64 | 0.082 | -69.01 | 0.00 |
| 2nd quintile | 0.33 | 0.02 | 15.61 | 0.00 |
| 3rd quintile | 0.23 | 0.02 | 10.65 | 0.00 |
| 4th quintile | 0.01 | 0.02 | 0.32 | 0.75 |
| 5th quintile | 0.93 | 0.02 | 39.01 | 0.00 |

**Table S6.** The stratified quasi-Poisson analysis. Results from stratified quasi-Poisson regression analysis. Counties were divided into strata based on their propensity scores quintile. Then, we fit a quasi-Poisson regression in each stratum using the number of COVID-19 deaths as the outcome and influenza vaccination coverage as the independent variable after adjusting for the propensity score. Shown is the intercept and coefficient for vaccination coverage from the quasi-Poisson model in each stratum.

| Variable | estimate | std.error | t-statistic | p.value |
| --- | --- | --- | --- | --- |
| 1st Quintile Intercept | -8.55 | 0.37 | -22.93 | 0.00 |
| 1st Quintile Immunization | -0.07 | 1.05 | -0.07 | 0.949 |
| 2nd Quintile Intercept | -9.43 | 0.58 | -16.29 | 0.00 |
| 2nd Quintile Immunization | 2.89 | 1.41 | 2.05 | 0.04 |
| 3rd Quintile Intercept | -8.31 | 0.77 | -10.85 | 0.00 |
| 3rd Quintile Immunization | -0.50 | 1.78 | -0.28 | 0.78 |
| 4th Quintile Intercept | -6.44 | 0.72 | -8.92 | 0.00 |
| 4th Quintile Immunization | -4.63 | 1.56 | -2.97 | 0.00 |
| 5th Quintile Intercept | -4.80 | 1.43 | -3.34 | 0.00 |
| 5th Quintile Immunization | -5.07 | 2.81 | -1.80 | 0.07 |

**Table S7.**
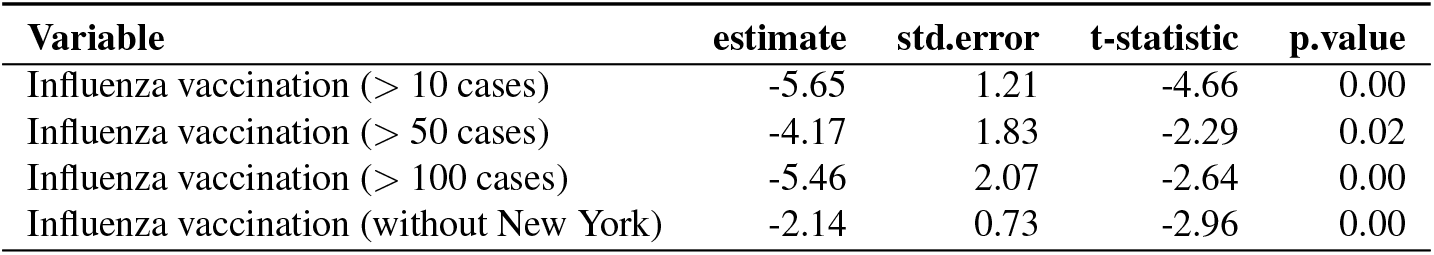
Effects of vaccination coverage using propensity score as a continuous variable. The table summarized the results from distinct quasi-Poisson regressions in which the number of counties included varies based on the minimum number of cases, or in which New York State counties were excluded. In each model, the number of COVID-19 deaths was used as the outcome and influenza vaccination coverage in people >= 65 together with propensity score (continuous) were used as the independent variables. Total population was used as offset.

**Table S8.** The effect of influenza vaccination on COVID-19 mortality in the U.S directly adjusting for the confounding variables. We clustered the 42 candidate variables based on their correlation with each other, then selected only one variable from each cluster. The selected variables were used as linear terms in the quasi-Poisson model.

| Variable | estimate | std.error | t-statistic | p.value |
| --- | --- | --- | --- | --- |
| (Intercept) | 31.16 | 6.19 | 5.04 | 0.00 |
| Influenza vaccination in people $\geq 65$ years | -4.67 | 0.77 | -6.07 | 0.00 |
| Family households | -3.40 | 0.70 | -4.85 | 0.00 |
| Ratio of hospital beds (per 1000 people) | 13.78 | 20.26 | 0.68 | 0.50 |
| Asthma | 15.50 | 4.42 | 3.50 | 0.00 |
| Breast cancer | 58.96 | 7.65 | 7.71 | 0.00 |
| Lung cancer | -17.92 | 16.66 | -1.08 | 0.28 |
| Chronic obstructive pulmonary disease | 5.15 | 2.21 | 2.33 | 0.02 |
| Hypertension | 0.90 | 0.98 | 0.92 | 0.36 |
| Median income | 0.00 | 0.00 | 7.44 | 0.00 |
| Summer temperature | -0.02 | 0.02 | -0.66 | 0.51 |
| Summer humidity | 0.03 | 0.01 | 3.66 | 0.00 |
| Winter temperature | -0.09 | 0.01 | -9.22 | 0.00 |
| Winter humidity | -0.08 | 0.01 | -9.01 | 0.00 |
| Age $\geq 65$ | 4.38 | 1.34 | 3.28 | 0.00 |
| Sex ratio | -0.07 | 0.01 | -5.08 | 0.00 |
| Days since the first case | 0.00 | 0.00 | 1.35 | 0.18 |

